# Apoptotic cells for therapeutic use in cytokine storm associated with sepsis

**DOI:** 10.1101/2020.12.03.20242586

**Authors:** Peter Vernon van Heerden, Avraham Abutbul, Sigal Sviri, Eitan Zlotnik, Ahmad Nama, Sebastian Zimro, Raja el-Amore, Yehudit Shabat, Barak Reicher, Batla Falah, Dror Mevorach

**Author notes:** Corresponding author: Prof. Dror Mevorach, Rheumatology and Rare Disease Research Center, The Wohl Institute for Translational Medicine, Hadassah-Hebrew University Medical Center and School, Kiryat Hadassah P.O.B 12000, Jerusalem 9112001, Israel. **Conflict of interest**: PV van Heerden received honoraria from Enlivex Ltd as a consultant. D Mevorach is the founder, and CMO of Enlivex Therapeutics Ltd.; Y Shabat and B Reicher are part of the research team of Enlivex Therapeutics Ltd. Enlivex Therapeutics administration was not involved in writing the manuscript.

## Abstract

**Background:** Sepsis has no proven specific pharmacologic treatment. Reported mortality in sepsis ranges from 30%–45%. This study was designed to determine the safety preliminary efficacy of allogenic apoptotic cells administered for immunomodulation in septic patients.

**Methods:** The primary aim of this phase IB study was to determine the safety profile of apoptotic cell infusion in subjects presenting to the emergency room with sepsis. Sepsis was determined by clinical infections and Sequential Organ Failure Assessment (SOFA) scores ≥2. The secondary aims were to measure organ dysfunction, intensive care unit (ICU) and hospital stays, and mortality, that were compared to historical controls. Exploratory endpoints included measuring immune modulator agents to elucidate the mechanism of action using Luminex® analysis.

**Findings:** Ten patients were treated with apoptotic cells, administered as a single dose or two sequential doses. All 10 patients had mild-to-moderate sepsis with a SOFA score range of 2–6 upon entering the study. No serious adverse events (SAEs) and no related AEs were reported. All 10 study subjects survived while matched historical controls had a mortality rate of 27%. The study subjects exhibited rapid resolution of organ dysfunction and had significantly shorter ICU lengths of stay compared to matched historical controls (p<0·0001). All patients had both elevated pro- and anti-inflammatory cytokines, chemokines and additional immune modulators that gradually decreased following treatment.

**Interpretation:** Administration of apoptotic cells to patients with mild-to-moderate sepsis was safe and had a significant immuno-modulating effect, leading to early resolution of the cytokine storm.

**Trial registration:** ClinicalTrials.gov Identifier: NCT03925857

**Funding:** The study was sponsored by Enlivex Therapeutics Ltd.

## Introduction

Sepsis, identified by the World Health Organization (WHO) as a global health priority, has no specific proven pharmacologic treatment other than appropriate antibiotics, intravenous fluids, vasopressors as needed, and possibly corticosteroids.^1-4^ The reported death rate from sepsis in hospitalized patients ranges between 30% and 45%.^5-10^

In sepsis, binding of either pathogen-associated molecular patterns (PAMPs) or damage-associated molecular patterns (DAMPs) to complement, toll-like receptors (TLRs), nucleotide-binding oligomerization domain (NOD)like receptors, retinoic acid-inducible gene (RIG)like receptors, mannose-binding lectin, and scavenger receptors, among others, induces a complex intracellular signaling system with redundant and complementary activities.^11,12^ Triggering of innate immunity assures a common response pattern, regulated by the level and variation of the repertoire of PAMPs and DAMPs, and the resulting signaling pathways that are activated. This sequence of events leads to the expression of several common classes of genes that are involved in inflammation, adaptive immunity, and cellular metabolism. The complementary nature of the pathways explains the overlapping yet unique early inflammatory response to common Gram-negative and Gram-positive bacterial, fungal, and viral infections, as well as tissue injury.

Interestingly, and as summarized recently, apoptotic cells in general, and Allocetra-OTS (Enlivex Therapeutics Ltd, Nes-Ziona, Israel) in particular, were shown to have a modulating effect on cytokine storms, with downregulation of both anti- and pro-inflammatory cytokines derived from PAMPs and DAMPs, in both animal and human *in vitro* models.^13^ Therefore, the current study was designed to examine the safety and the possible beneficial immuno-modulating effects of apoptotic cells (Allocetra-OTS, Enlilvex Therapeutics Ltd., Nes-Ziona, Israel) administered to patients presenting with sepsis. In a previous dose-escalating clinical study enrolling patients undergoing bone marrow transplantation who had an elevated cytokine profile, matched apoptotic cells were shown to be safe and efficacious with a dose-dependent effect starting at 140×10^6^ cells/kg.^14^ This dose, administered in either one or two treatments, was chosen for the current study. In addition, the outcome of study subjects in this safety trial were compared to historical matched controls to compare outcomes at 28 days.

## Methods

### Study design

The primary aim of this phase Ib study (ClinicalTrials.gov Identifier: NCT03925857) was to determine the safety profile and tolerability (dose-limiting toxicity) of Allocetra-OTS infusion in subjects presenting to the emergency room with sepsis. The secondary aims were to determine the preliminary efficacy on reducing organ dysfunction; intensive care unit (ICU), intermediate care unit (IMU) and hospital stays; and mortality.

Adult males and females aged 18–85 years, weighing at least 40 kg and with a life expectancy of at least 6 months at the time of the screening, who had a SOFA score >2 above baseline and had sepsis from presumed infection were included. Exclusion criteria included pregnancy, positive serology for HIV, performance status less than 80%, or serious organ dysfunction (e.g. left ventricular ejection fraction <40%, pulmonary forced vital capacity <60% of predicted, liver transaminases >2·5× the upper limit of normal, serum bilirubin >3 mg/dL, or creatinine >2·5 mg/dL.

Sequential Organ Failure Assessment (SOFA) scores were measured at enrollment and at each study time point. We also obtained blood samples for exploratory biological tests including pro- and anti-inflammatory cytokines, chemokines, growth factors, leptin, ghrelin, neutrophil gelatinase-associated lipocalin (NGAL), Triggering Receptor Expressed on Myeloid Cells 1 (TREM1), an endocrinology panel, cortisol, ACTH, FT3, FT4, TSH, growth hormone, glucagon, and insulin. An autoimmune serology panel that included ANA, anti-DNA, anti-RNP, anti-SSA, anti-SSB, cardiolipin IgG, and cardiolipin IgM was also taken.

Allocetra-OTS was administered as a single dose (cohort 1) of 140×10^6^ cells/kg on day +1 (day 0 was time of diagnosis at the ER) or in two doses (cohort 2) of 140×10^6^ cells/kg on days +1 and +3, following initiation of standard-of-care (SOC) treatment for septic patients, as outlined by the Surviving Sepsis Campaign.^14,15^ Interim safety analyses were performed by the Data and Safety Monitoring Board (DSMB) after three and six patients from cohort 1 had completed study day 14, and after four additional patients from cohort 2 had completed study day 28.

### Alloctera-OTS preparation

Enlivex Therapeutics Ltd. has developed a product named Allocetra-OTS based on the known activity of naturally occurring apoptotic cells to induce a pro-homeostatic state for both macrophages and dendritic cells (DCs)^13,14,16-18^ that contributes to maintenance of peripheral homeostasis of almost all immune-triggered mechanisms in sepsis. Allocetra-OTS is composed of irradiated non-HLA-matched mononuclear enriched leukocytes containing at least 40% early apoptotic cells, in the form of a liquid suspension that is administered intravenously (IV).

### Matched historical controls

The local ethics committee also approved (#0267-19-HMO) a search for matched controls that were selected from the digital records of all hospitalized patients treated in the same units as the study patients. Controls were selected from all patients hospitalized in Hadassah Medical Center at IMU or ICU with diagnosis of sepsis originated from pneumonia, UTI, Biliary infection or endovascular infection. The total number was 24,172 patients including 18,078 that were pneumonia patients, 5370 UTI patients, 629 patients with biliary tract infection and 95 patients with endovascular infection. Matching the characteristics of each of the 10 sepsis patients that were treated with Allocetra was done with full match to source of infection, SOFA score ±2, age ±7 years, and gender, resulted in 37 matched historical control patients, of whom 19 were with pneumonia (Table 1). The process of identifying the suitable patients was done by two persons and reviewed by a third. All data was found in patients’ charts and SOFA score was calculated retrospectively. Outcomes for the matched controls were compared to those of the study cohorts at 28 days. Source data was verified to confirm that the reported Qualified Data provided in the report match the source.

**Table 1.** Characteristics Patients and Historical Matched-Controls.

|  | **Total Sepsis Treated patients**<br>**(n = 10, 100%)** | **Total Sepsis Matched-Controls**<br>**(n = 37, 100%)** | **Sepsis source: Pneumonia Treated Patients**<br>**(n=5, 50%)** | **Sepsis source: Pneumonia Matched Controls (n=19, 51.3%)** |  |
| --- | --- | --- | --- | --- | --- |
| Mean age (range) | 71.5 (51–83) | 73.1 (50-79) | 67.8 (51–79) | 67.8 (50–79) |  |
| Male/female | 8/2 | 31/6 | 4/1 | 15/4 |  |
| Mortality | 0/10 | 10/37 (27%) | 0/5 | 6/19 (31.5%) |  |
| **Patient** | **SOFA score at screening** | **Predicted mortality (%) based on SOFA at presentation** | **APACHE II Score** | **Predicted mortality % based on APACHE II** | **28-day outcome in patients treated with Allocetra-OTS** |
| 01-001<br>Pneumonia | 2 | 8 | 12 | 15.9 | Alive. Sepsis and organ dysfunction resolved |
| 01-002<br>Pneumonia | 3 | 10 | 9 | 9.9 | Alive. Sepsis and organ dysfunction resolved |
| 01-003<br>Pneumonia | 6 | 16 | 15 | 21 | Alive. Sepsis and organ dysfunction resolved |
| 01-006<br>Pneumonia | 3 | 10 | 11 | 12.9 | Alive. Sepsis and organ dysfunction resolved |
| 01-007<br>Endovascular | 6 | 16 | 21 | 38.9 | Alive. Sepsis and organ dysfunction resolved |
| 01-008<br>UTI | 3 | 10 | 9 | 9.9 | Alive. Sepsis and organ dysfunction resolved |
| 01-009<br>Biliary | 3 | 10 | 13 | 18.6 | Alive. Sepsis and organ dysfunction resolved |
| 01-010<br>Biliary | 5 | 13 | 16 | 23.0 | Alive. Sepsis and organ dysfunction resolved |
| 01-011<br>Biliary | 2 | 8 | 8 | 8.9 | Alive. Sepsis and organ dysfunction resolved |
| 01-012<br>Pneumonia | 2 | 8 | 15 | 21 | Alive. Sepsis and organ dysfunction resolved |
| Average (range) | 3.4 (2–6) | 11.9 (8–16)% | 12.3 (8–21) | 16.8 (9.9–38.9)% |  |

### Luminex® Cytokine/ Chemokine analysis

Serum cytokine/chemokine measurement was performed using the Luminex MAGPIX system (Luminex Corp, Texas, USA) and analyzed with Milliplex analysis software (Millipore MA, USA).

### ELISA analysis

The following cytokines/ chemokines were measured by sandwich ELISA kits: IL-18 (R&D), MCP-3 (R&D), TNFR1 (R&D), TREM-1 (R&D), procalcitonin (PCT) (IBL-America, MN, USA).

### Autoantibodies

and anti-HLA antibodies were analyzed by the central laboratory of the Hadassah-Hebrew University Medical Center.

### Statistics. Clinical data

Mortality was analyzed as time-to-event measurement and comparisons between treatment arms were presented using Kaplan Meier curves and the log rank test. Expected mortality estimation was based on patients’ Acute Physiology and Chronic Health Evaluation (APACHE) II scores.^19^ Changes in SOFA score from entry into research to day 5, 7 and 28, maximum increase in SOFA score, duration of hospitalization and ICU/IMU stays were analyzed as continuous variables. Means, standard deviations, medians, interquartile ranges, and ranges in each treatment group were presented and compared using the t-test or Wilcoxon rank sum test, as appropriate. Area under the curve for change in SOFA score was calculated using the trapezoidal rule and also analyzed as a continuous variable. Duration of hospitalization and ICU/IMU stays were analyzed as time to first discharge, presenting the Kaplan-Meier curves and comparing treatment groups using the log rank test. All analyses were repeated for the entire population as well as for subgroups receiving one or two Allocetra-OTS doses, and for pneumonia alone. Statistical analysis was performed using R (R Statistical Data Analysis).

### Laboratory and exploratory tests

Correlation of any parameter to clinical score was evaluated by the Pearson correlation or by Spearman’s rank correlation coefficient, with a coefficient >0·7 or <-0·7 considered a strong correlation. Statistical analyses for laboratory tests were performed using GraphPad (Prism, San Diego, CA, USA).

### Study approval

The study was approved by the local ethics committee and the Israeli Ministry of Health and was performed according to institutional guidelines. Written informed consent was obtained in accordance with the Declaration of Helsinki from all study subjects and apoptotic cell donors.

## Results

Twelve patients were screened, and ten were included in the study; patients 04 and 05 did not meet inclusion criteria. The DSMB met following inclusion of three (patients 01–03) and six patients (patients 01–03 and 06–08) for a safety review and approval to continue the study, and for final review after 10 patients. Allocetra-OTS infusion in the first three patients met the protocol for defined safety criteria at day 14 and the study proceeded to the second round of patient recruitment (patients 06–08) at the same dose, which also met the protocol-defined criteria for safety. The study then proceeded to recruit four additional patients (cohort 2, patients 09–12), who received two doses of Allocetra-OTS. Patient characteristics are presented in Table 1.

### Clinical course

All study subjects fitted the definition of sepsis and inclusion criteria and were hospitalized on admission in either the ICU or the IMU. All patients had a Glasgow Coma Score (GCS) of at least 13 (verbal 5/5) at enrollment due to the requirement to obtain consent directly from the study subject. At enrollment, the average APACHE II score was 12·9 (range 8–21) and the average SOFA score upon entry into the study was 3·4 (range 2–6). All patients fulfilled inclusion and exclusion criteria and met the 2016 definition of sepsis.^20^ In this study, any source of infection was included, and the patients presented with four types of infections: pneumonia (five patients), biliary tract infection (three patients), endovascular (one), and urinary tract infection (one).

All patients had at least two organ systems involved (range 2–5 systems). Acute kidney injury (three patients), cardiovascular involvement (three patients), hepatic involvement (seven patients), hematological (eight patients), and pulmonary involvement (five patients) were seen in the treated patients before investigational product (IP) administration. All patients recovered from the septic condition and were discharged alive from the hospital following the resolution of sepsis and were alive at completion of 28 days of follow-up.

### Laboratory results

Laboratory evaluation included complete blood count (CBC), biochemistry, blood gases, C-reactive protein (CRP), and lactate. Figure 1 shows their levels during the study period. In 5/10 patients, elevated white blood cells (WBC) counts were evident in the first days, with gradual return to normal levels (Figure 1A). Neutrophilia was observed on admission in 6/10 patients (Figure 1B) and lymphopenia in 9/10 patients (Figure 1C). All patients had a gradual increase in lymphocyte numbers and 6/9 (66%) recovered to normal levels whereas 3/9 (33%, patients 03, 06, 07) had moderate recovery levels. Elevated CRP declined in parallel with resolution of inflammation (Figure 1D). Two patients (07 and 12) had a slower decline of CRP. The first (patient 07) was a patient on chronic dialysis presenting with an endovascular infection that needed antibiotics for 6 weeks, and the second (12) was a patient with pneumonia and pleural effusion. Lactate levels were elevated in 3/10 patients upon admission (01, 02, 08) and were normal or near normal in the following days. Lactate levels were elevated in 3/10 patients upon admission (01, 02, 08) and were normal or near normal in the following days. Three patients had higher lactate levels on day 28 (01, 03, 09) despite clinical and laboratory resolution of sepsis, in the range of 3·4 to 6·5 mmol/L (normal value up to 2·2 mmol/L).

**Figure 1.**
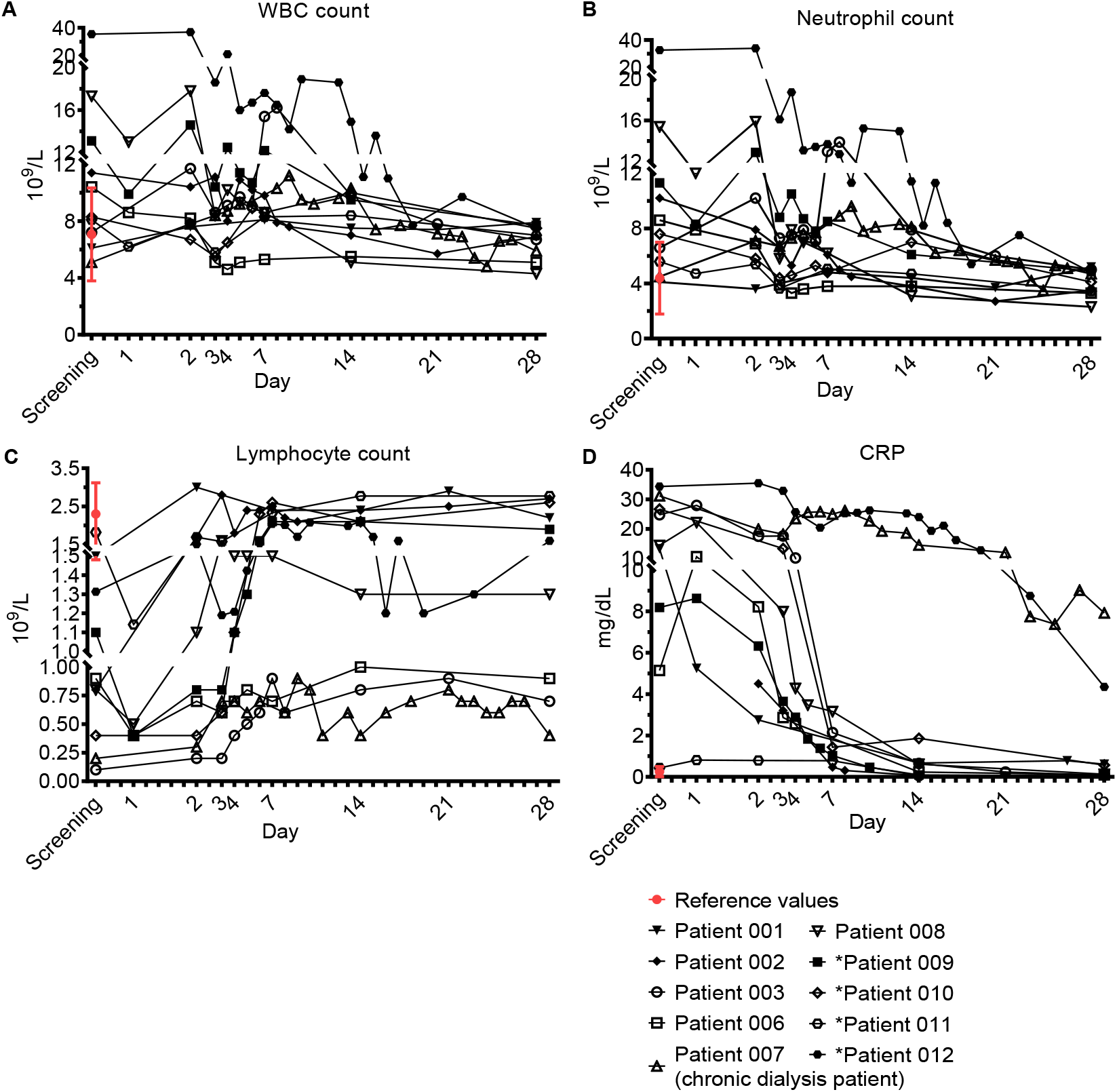
Acute phase markers. Complete blood count (CBC) and C-reactive protein (CRP). Blood counts of fresh peripheral patient blood ethylenediamine tetraacetic acid (EDTA) samples were performed using blood analyzer. The differential of white blood cells (WBC), i.e. WBC subpopulations, was calculated. ‘Screening’ is a time point with a blood count that was performed 24±6h before treatment with Allocetra-OTS, to test for the patient’s eligibility for Allocetra-OTS treatment. ‘Day 1’ is a blood sample just prior to Allocetra-OTS infusion. Numbers (10^9^ cells/L) of (**A**) WBCs, (**B**) neutrophils, and (**C**) lymphocytes, are shown. (**D**) CRP levels of all patients in the indicated time points. **\***Patients 09–12 received two doses of Allocetra-OTS (days 1 and 3).

### Safety parameters

Safety was evaluated by serious adverse events (SAEs) and adverse events (AEs). All patients survived 28 days of follow-up. Of note, efficacy parameters (presented below), like survival, overlapped with additional safety parameters. There were no serious unexpected serious adverse reactions (SUSARs) and none of the subjects experienced an SAE, severe or moderate AE, or discontinued the study due to an AE. Nearly all subjects (9/10; 90%) experienced at least 1 AE and on average 4.4 AEs; The most common AEs were associated with laboratory investigations (Table 2) and all were mild in intensity and most were unrelated to IP. The six possibly related AEs were transiently elevated temperature, mild transient tachycardia, transient hypoglycemia, and transient dizziness, each in one patient, and rigor episodes in two patients receiving one dose. All these AEs could have been related to the septic condition. However, since the two episodes of rigor occurred in patients receiving a high fusion rate (>150 mL/h), the last 8 IP administrations were given at a slower rate (up to 108 mL/hr) with no rigors documented. Autoantibody induction, including ANA, anticardiolipin, and anti-DNA were examined during the study period and found to be negative.

**Table 2.**
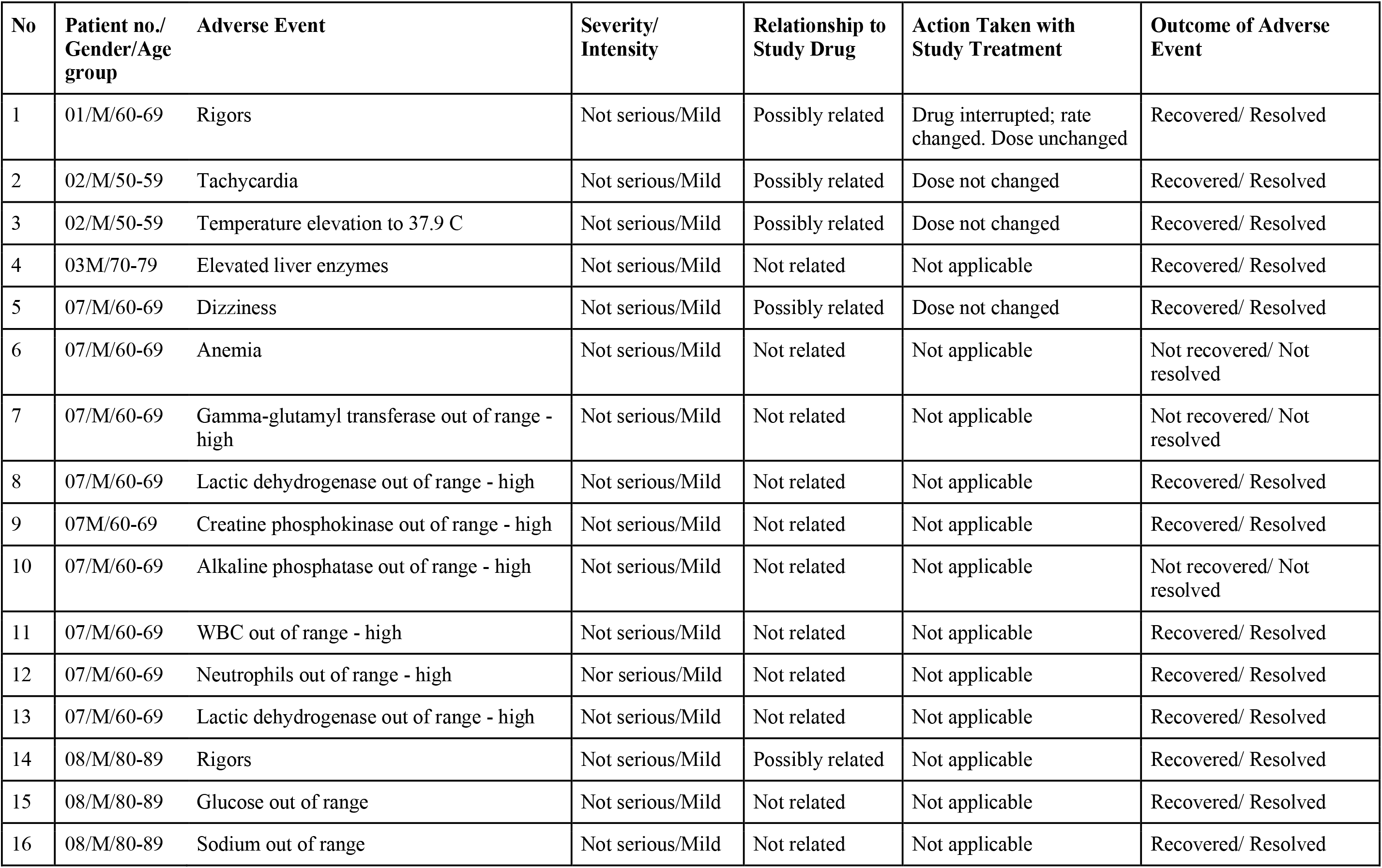
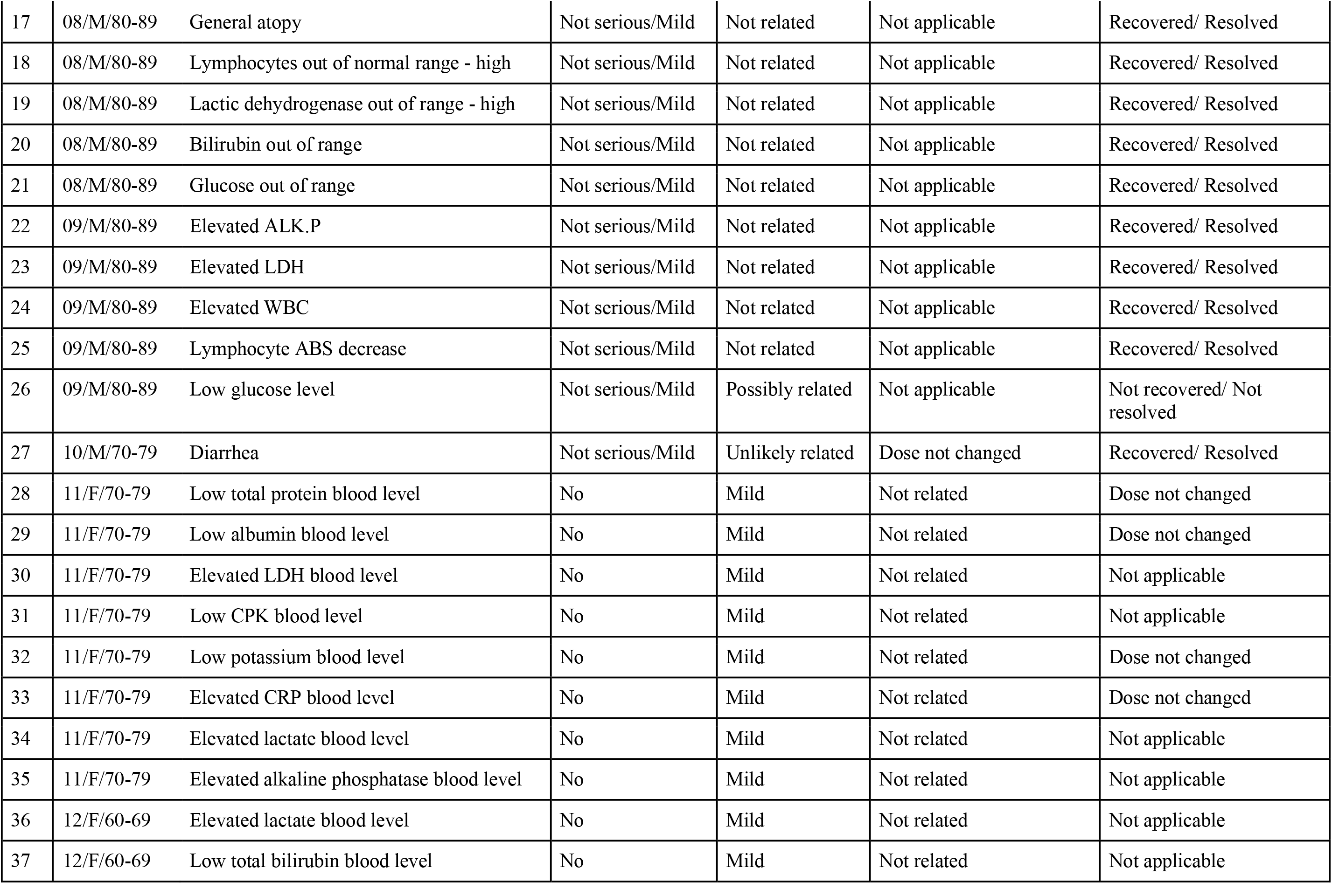
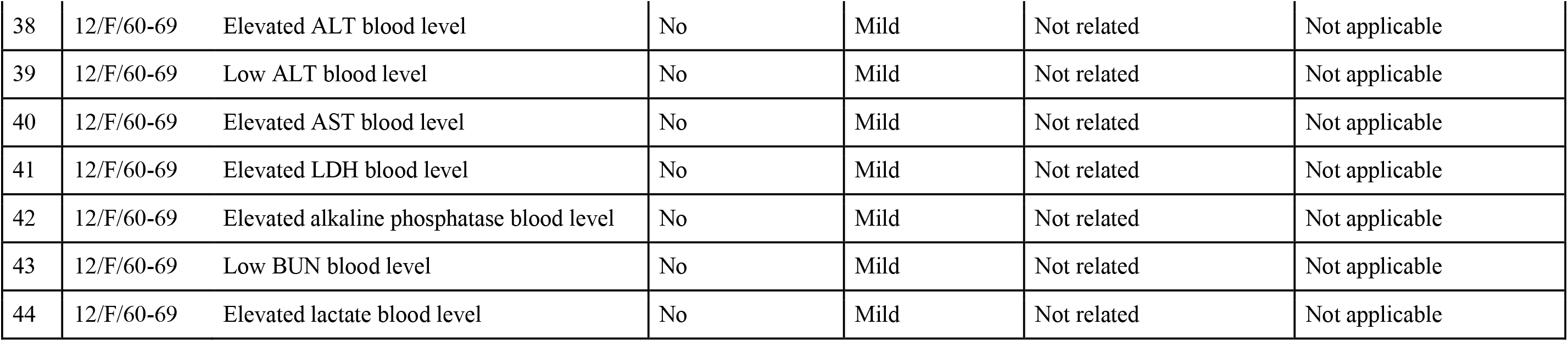
Complete list of Adverse Events.

### Autoimmunity, anti-HLA antibodies, and autoantibodies

No anti-HLA antibodies developed within 28 days following one dosage in the first 6 patients. Autoantibodies and autoimmunity were evaluated at initial screening and on day 28. No autoimmunity developed in any of the study subjects during the study period. The antinuclear antibody screen (ANA), which is considered a major screening tool for autoantibodies, was negative at presentation in 8/10 patients and remained negative in all eight at day 28. In two patients it was low-positive before IP administration; it remained low-positive in one and disappeared in the second. IgM and IgG anticardiolipin were negative in 10/10 patients at screening and remained negative in all.

Anti-DNA was not examined at initial screening, due to negative ANA in most patients, but was negative in all patients examined at day 28. Anti-SSA/SSB/RNP/Sm were examined mainly on day 28 and in some patients at initial screening. Only one patient (08) had very low positive RNP (20·61 vs 20, which is defined as negative) on day 28. This was considered non-significant.

In conclusion, after administration of 14 IV doses of Allocetra-OTS in 10 patients, there was no evidence of autoimmunity or autoantibodies.

### Preliminary efficacy results: mortality

No deaths occurred among the 10 study subjects. APACHE II scores for subjects in the current study are presented in Table 1. The average score at diagnosis was 12·9 (range 8–21). Overall probability of mortality was 16·8% (range 9·9–38·9%), with no significant differences between the subgroups of patients.

In patients with sepsis, an elevated SOFA score at presentation also reflects an increased risk of mortality. In one study that included >180,000 patients hospitalized with sepsis, a SOFA score of 2– 6 at presentation (Table 1) corresponded to a predicted mortality of 8–16%.^21^

In addition, a total of 37 matched controls were identified for the 10 patients in the study based on the criteria of sepsis, admission to ICU/IMU, source of infection, SOFA score ±2, age ±7 years, and gender. Table 1 shows the characteristics of controls compared to study subjects. A comparison of survival between the study group and historical controls, in all the patients and in the subgroup of patients with pneumonia is shown in Figure 2. Among the 37 matched historical control patients, 10 died (27%, p-value using log rank test=0·078). Although a bigger sample is needed to draw definite statistical conclusions regarding survival, predicted mortality based on the APACHE II and SOFA scores at presentation would have been a high probability of at least one death in these 10 patients (85%).

**Figure 2.**
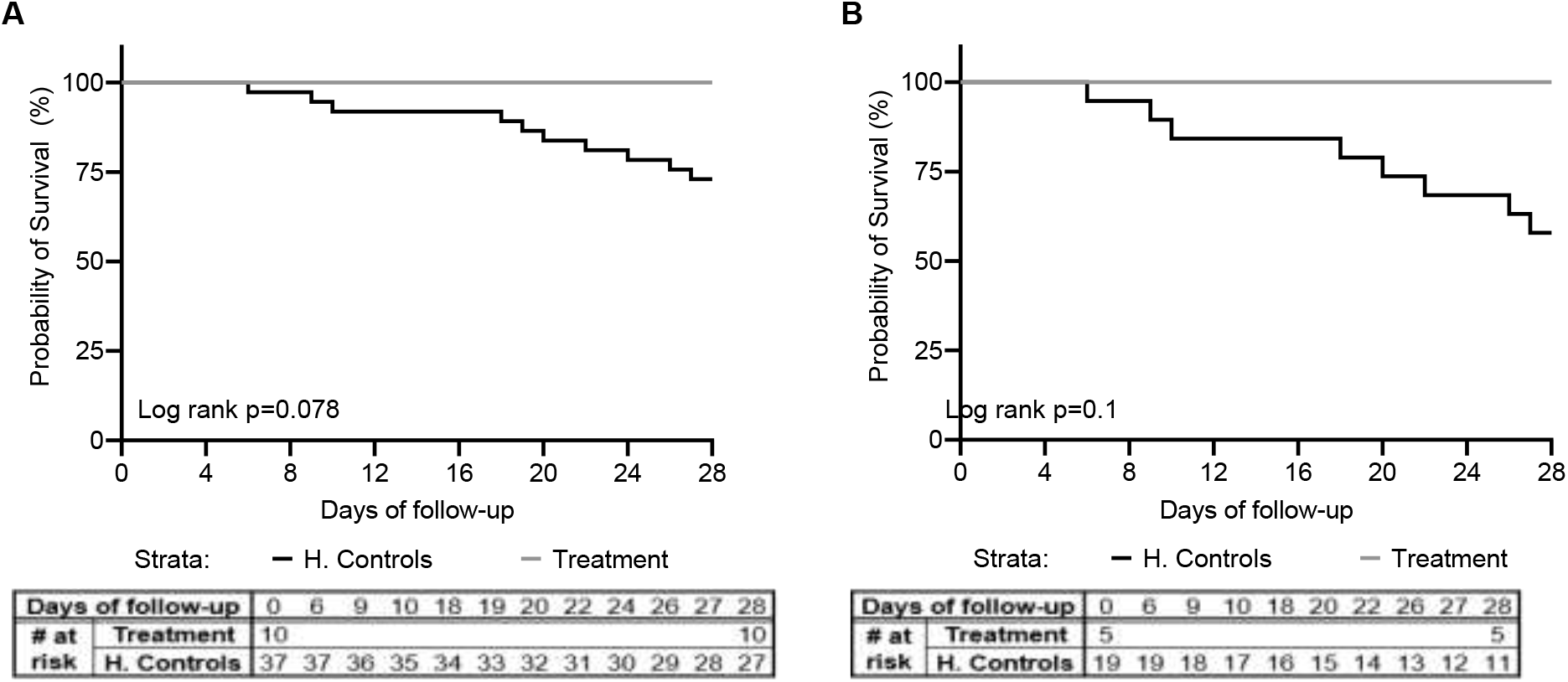
Kaplan-Meier survival curve. A comparison of survival between the study group (*n*=10) and matched-historical controls from the same hospital (*n*=37) for **(A)** all patients, and **(B)** patients with pneumonia (*n*=5 for the study group, *n*=19 for the historical controls), is shown. Matched controls were selected from all patients admitted to ICU under the diagnosis of sepsis between 2014–2019 and were matched by gender, age±7, Sequential Organ Failure Assessment (SOFA) score ±2 at presentation, and source of infection.

### Preliminary efficacy results: mortality

No deaths occurred among the 10 study subjects. The expected number of deaths in hospitalized septic patients ranges between 30% and 45%;^5,6,8-10^ however, in the specific studied population, with a GCS of at least 13 it could significantly lower. This should be taken into account in the discussion below.

The APACHE II scoring system has been shown to be an accurate measurement of patient severity and correlates strongly with outcome in critically ill patients.^16,18^ APACHE II is measured during the first 24 h of ICU admission; the maximum score is 71. Patients with a score of 25 have a 50% predicted mortality rate, and those with a score over 35 have a predicted mortality rate of 80%. APACHE II scores for subjects in the current study are presented in Table 1. The average score at diagnosis was 12·9 (range 8–21). The probability of survival for the 10 individual patients based on their APACHE II scores was calculated.^21^ Overall probability of mortality was 16·8% (range 8·9– 38·9%), with no significant differences between the subgroups of patients who received one or two doses, between the five pneumonia patients and three patients with biliary infection, or the two women versus the eight men. Current state-of-the-art pre-, intra-, and post-ICU treatment protocols for emergency admissions with sepsis may have improved physiological values and corrected physiological abnormalities, resulting in lower mortality compared to the rates predicted by the APACHE II scores. However, it should be noted that each of the 10 patients enrolled in our study was in real danger of mortality based on their admission APACHE II scores. The estimated probability that at least one study subject would die was 85%.

In patients with sepsis, an elevated SOFA score at presentation also reflects an increased risk of mortality. In one study that included >180,000 patients hospitalized with sepsis, a SOFA score of 2– 6 at presentation (Table 1) corresponded to a predicted mortality of 8–16%.^21^

### Organ dysfunction

Organ dysfunction improvement was a preliminary efficacy parameter tested in this study. No residual organ dysfunction was seen in any of the 10 study subjects at 28 days. **CNS**. All patients finished 28 days of follow-up with a GCS of 15/15. **Kidneys**. Apart from patient 07, who was on chronic dialysis at admission, 3/9 patients (33%) had new-onset renal injury, and all had completely recovered to baseline kidney function as measured by creatinine level at 28 days. **Lungs**. 5/10 (50%) of patients had lung involvement. No patient required mechanical ventilation. All patients recovered from lung dysfunction, had normal oxygen saturation, and needed no oxygen supplement at discharge. **Cardiovascular**. Three patients had mean arterial pressure <70mmHg but none needed vasopressors. Cardiac evaluation was made using clinical evaluation, ECG in all patients, troponin if indicated, and echocardiogram as indicated (needed in one patient). All patients had normal sinus rhythm or tachycardia upon screening, with no evidence of ischemia during their illness. One patient with known paroxysmal atrial fibrillation had a transient episode of atrial fibrillation. Troponin was measured in four patients and was normal in all. One patient (01) underwent transthoracic echocardiography during his admission for the investigation of chest pain and elevated troponin levels following an episode of supraventricular tachycardia and electrical shock (DC cardioversion) before screening and entry to the study. Follow-up echocardiography in this patient was performed at day 28. The results were comparable to the first evaluation. **Hematological**. Significant thrombocytopenia occurred in 8/10 patients (80%) with complete recovery in all. **Liver**. Hyperbilirubinemia occurred in four patients (40%) and three patients had a biliary tract infection, all four had a complete recovery. Elevated liver enzymes (AST and ALT) >3 of normal range were seen in 5/10 patients with a complete recovery in all. In the absence of days on respirator and days on vasopressors, we used SOFA score to evaluate organ dysfunction. The SOFA score was introduced to describe organ failure severity in patients with sepsis, including a 4-point assessment of dysfunction in each of six organ systems.^17,19,22^ In patients with sepsis, a SOFA score ≥2 at presentation reflects clinically relevant organ dysfunction and an increased risk of adverse outcomes.^21^ Apart for acute organ injury, we measured changes in the SOFA score from just before IP administration, maximal SOFA score, AUC of SOFA score above baseline and time to SOFA score <2 and compared it to the matched-historical controls

As shown in Table 1 and Figure 3, patients had SOFA scores that correlate to mild to moderate sepsis (patients 02–06). However, despite the similarity to historical-matched controls of SOFA scores at entry (average of 3·4 versus 3·47), the enrollment SOFA score was the highest for the treated patients that did not progress further (0 points) following treatment, while it progressed with a mean maximal increase of 3·57 (range 0–15, median 1) in the historical control population (Figure 3A; p< <0·0001, t-test and Wilcoxon), suggesting inhibition of organ dysfunction deterioration with Allocetra-OTS treatment. In historical-matched controls with pneumonia, it went even higher, to a mean of +4·42 (range 015, median 4) (p<0·0001, t-test, and 0·0166, Wilcoxon).

**Figure 3.**
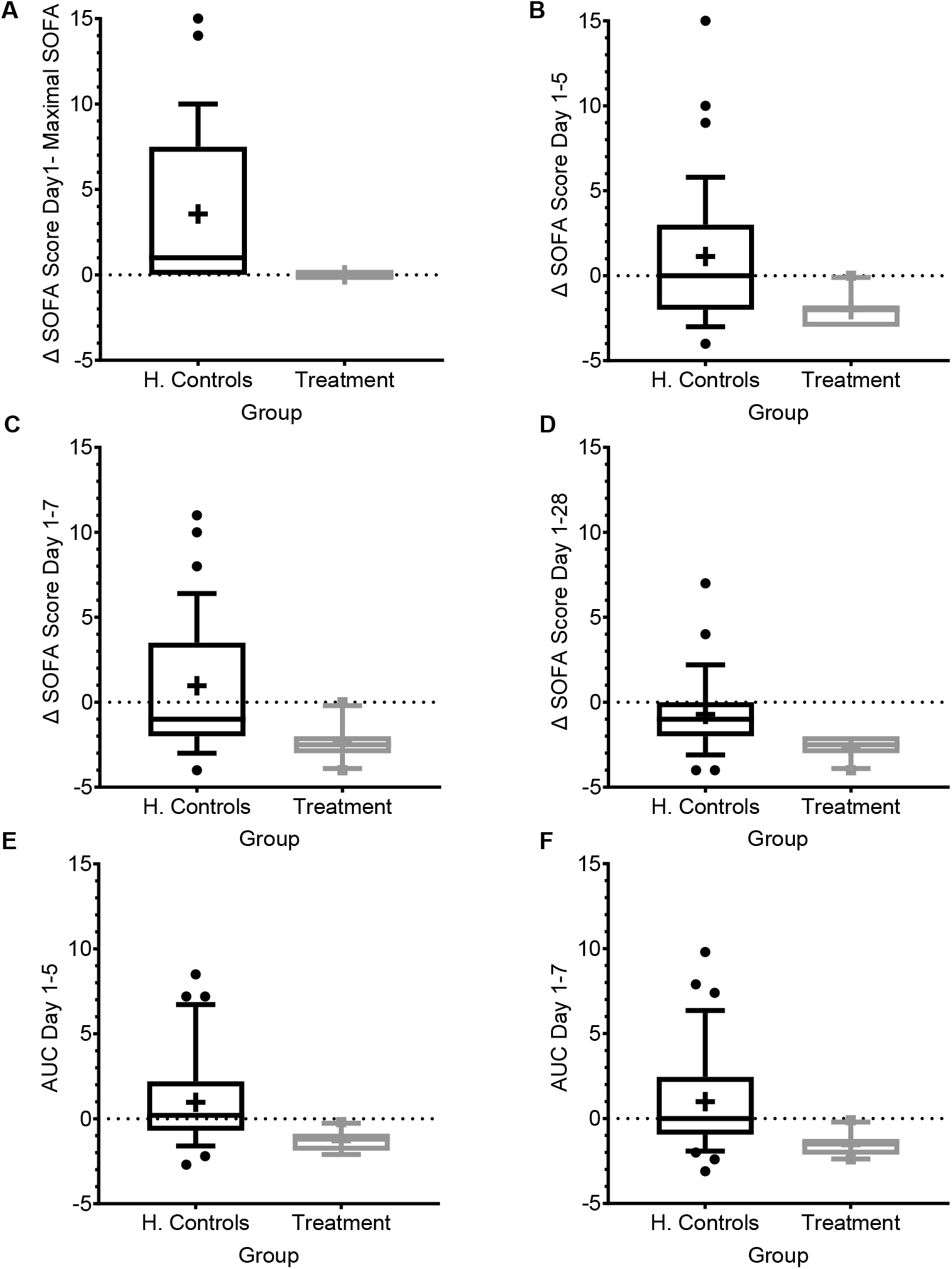
SOFA score progression during sepsis. **(A)** SOFA score at presentation compared to maximal SOFA score. **(B–D)** Change in SOFA score before administration of Allocetra-OTS and day 5 **(B)**, day 7 **(C)**, and day 28 **(D)**. Average delta area under the curve (AUC) in the first **(E)** five days, and **(F)** seven days. Matched controls were selected from all patients admitted to ICU under the diagnosis of sepsis between 2014–2019 and were matched by gender, age ±7 years, SOFA score ±2 at presentation, and source of infection. Data is presented as the median within the inter-quartile range (IQR); mean values are marked with a ‘+’ sign; error bars represent the 10-90 percentile, with outliers presented.

We further measured the change in mean SOFA score between day 1 before treatment with Allocetra-OTS and at days 5, 7, and 28, which was significantly higher in the historical-matched controls than the treated patients (Figure 3 B-D; average delta SOFA score between day 1–5, day 1–7, and day 1– 28, p<0·0001, t test for all, and 0·0035, 0·001, and 0·0019, Wilcoxon, respectively). We also measured the average change in the area under the curve (AUC) between days 1 and 5 and between days 1 and 7 (Figure 3 E,F); all found significantly ameliorated (average AUC between day 1–5 and day 1–7, both p<0·0001, t test, and 0·0011, Wilcoxon). In historical matched controls with pneumonia, it went even higher, to a mean increase of 4·42 (range 0–15, median 4; p<0·0001 t-test, and 0·0166 Wilcoxon). These data suggest that the favorable clinical outcomes are reflected by lack of SOFA score progression as manifested by maximal SOFA score, delta SOFA score, and the average change in AUC.

### Time in ICU/IMU and hospital

Since organ dysfunction was significantly improved and no mortality occurred, we were next interested to verify that these promising results were expressed in the duration of ICU/IMU and hospital stay. Since all patients in the study group survived, we analyzed duration of stay in the hospital and in ICU/IMU for all patients in the study and for all surviving patients in the matched historical control group (Figure 4).

**Figure 4.**
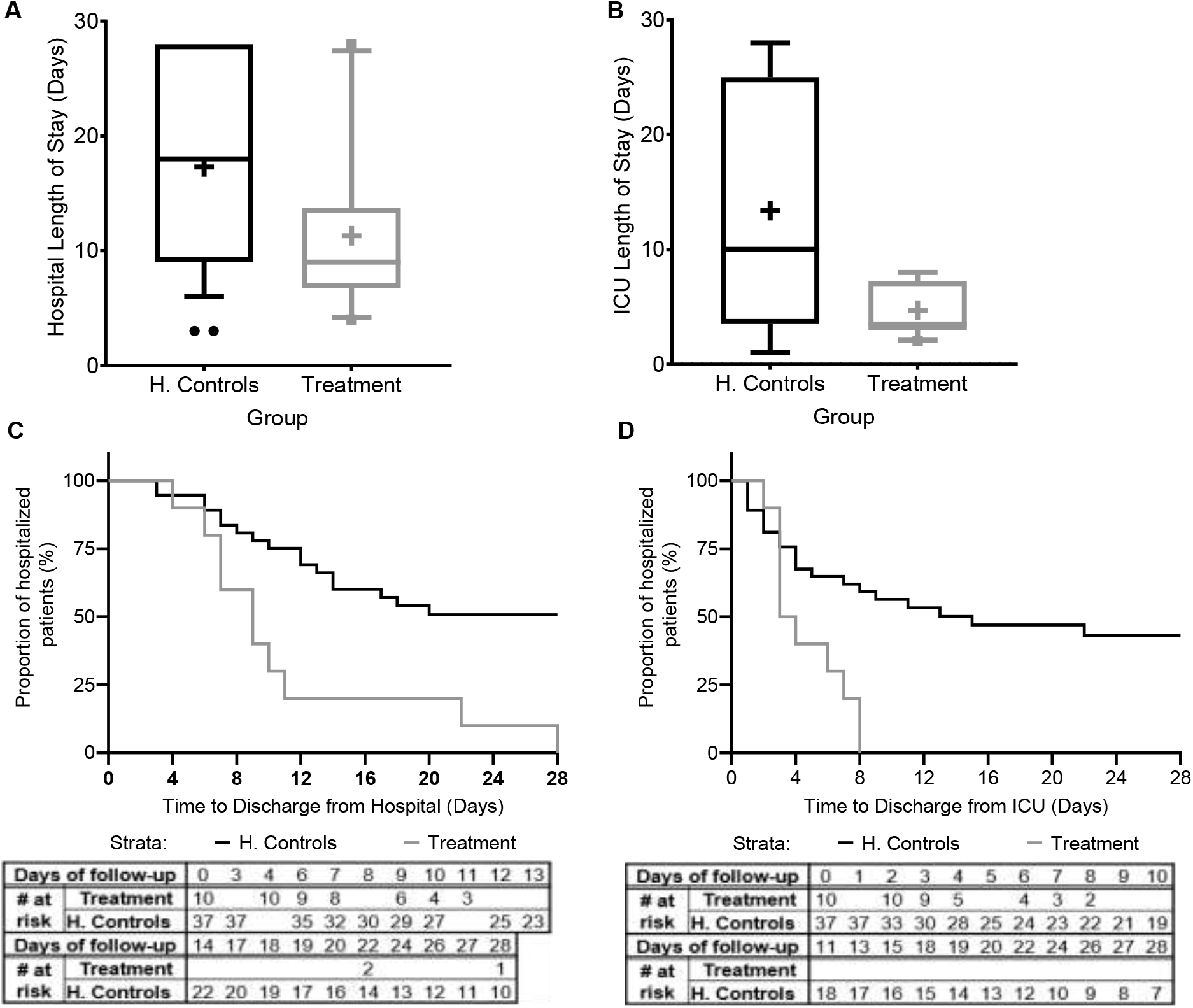
Hospital and ICU/IMU duration of stay. The duration of stay for all patients in the study and for all patients in the matched historical control group who survived is presented **(A)** in the hospital and **(B)** in the Intensive Care Unit (ICU) or the intermediate medical unit (IMU). Kaplan-Meier curves describing time-to-discharge from the **(C)** hospital and **(D)** ICU/IMU are presented. Matched controls were selected from all patients admitted to ICU/IMU with the diagnosis of sepsis between 2014–2019 and were matched by gender, age ±7, SOFA score ±2 at presentation, and source of infection. Data is presented as the median within the interquartile range (IQR); mean values are marked with a ‘+’ sign; error bars represent the 10-90 percentile, with outliers presented.

The mean hospital length-of-stay was significantly shorter for all sepsis patients treated with the Allocetra-OTS (11·4 ± 7·57 days, range 4–28, median 9), and for patients with pneumonia (11·2 ± 6·38 days, range 6–22, median 10), compared to the averages for matched-historical controls of for all patients (Figure 4A; 17·3 ± 8·8 days, 3–28, median 17; p< 0·0488 t-test, p<0·042 Wilcoxon), and for patients with pneumonia, average 18·84 days (range 6–28, median 19; p< 0·0552 t-test, p< 0·0484, Wilcoxon). The ICU length-of-stay was significantly shorter for all treated patients, average 4·7 days (range 2–8, median 4), and for patients with pneumonia, average 3·4 days (range 2–6, median 2) compared to averages for all matched controls, 11·1 days (Figure 4B; 1–28, median 8; p< <0·0001 t-test, p< 0·092, Wilcoxon) and patients with pneumonia, 13·89 days (range 1–28, median 11; p<0.0001 t-test, p< 0.0233 Wilcoxon).

In addition, time-to-discharge from the hospital and/or ICU/IMU (Figure 4 C,D) was compared. Length of hospitalization was analyzed as a time-to-event variable, comparing time to discharge from the general hospital and the ICU between treatment groups. In this analysis, events of mortality were referred to as no discharge event and the length of follow-up was censored at time of death. Times to hospital and ICU/IMU discharge were significantly shorter for the treated group (log rank p=0·00085 and p=0·00096, respectively).

### Exploratory endpoints. Effect of Allocetra-OTS on cytokines/chemokines/ growth factors and immuno-modulating agents. Pro-inflammatory cytokines

Pro-inflammatory cytokines regulate early responses to bacterial infection and mediate the early acute phase in sepsis. Cytokines like IL-1, IL-6, and TNF-α act as endogenous pyrogens, upregulate the synthesis of secondary mediators and other pro-inflammatory cytokines by both macrophages and mesenchymal cells such as fibroblasts, epithelial, and endothelial cells, and stimulate the production of acute-phase proteins or attract inflammatory cells.^23^ Eight pro-inflammatory cytokines were tested, including IL-6, TNF-α, IL-1β, IL-12p70, IL-18, IL-23, IFN-γ and IL-13; five of those were detectable (IL-6, TNF-α, IL-1β, IL, IL-18, IFN-γ) and are presented here (Figure 5A).

**Figure 5A.**
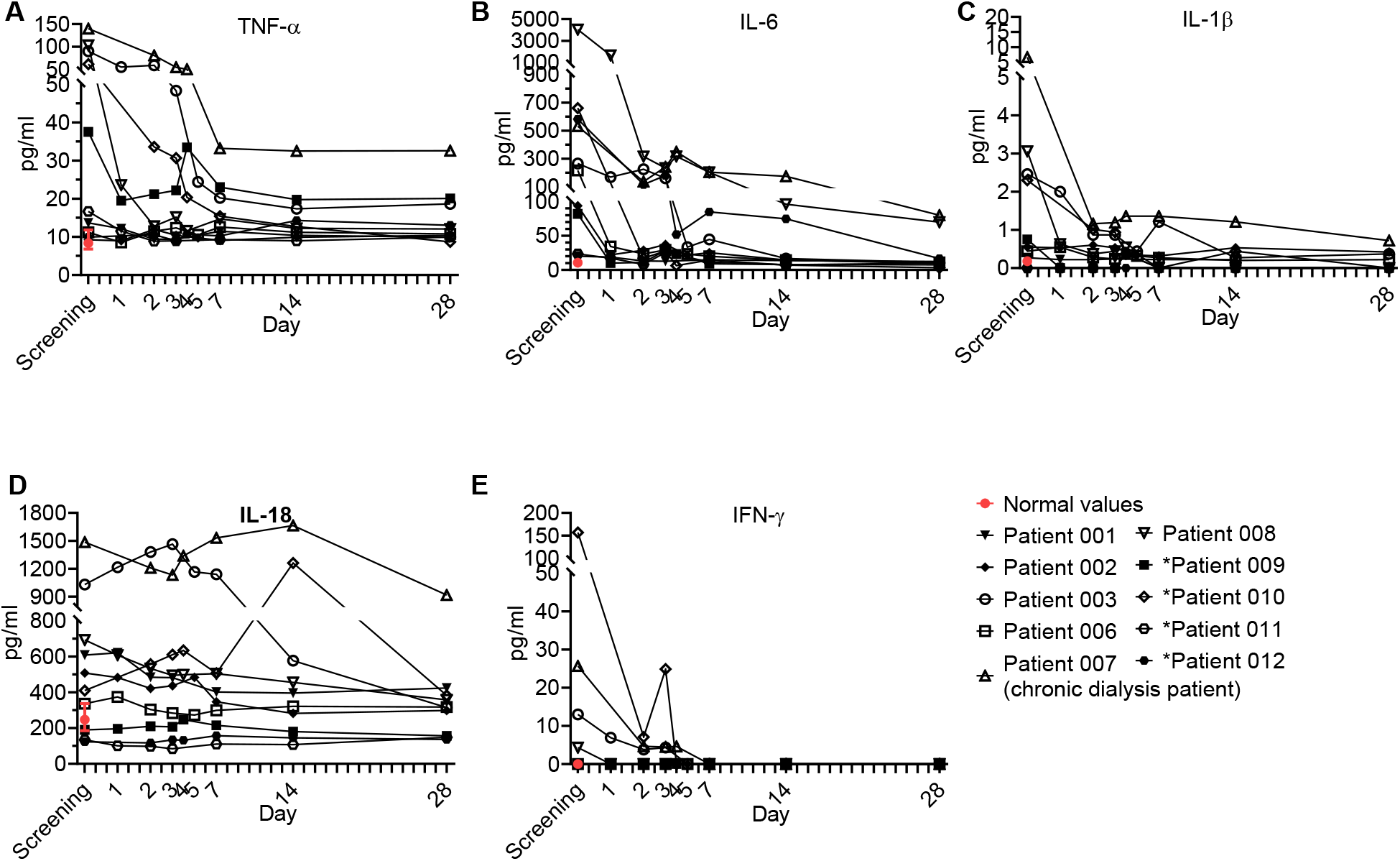
Pro-inflammatory cytokine kinetics during sepsis. Serum was obtained from patients at the indicated times and cytokine analysis was performed as described in Methods. The serum of three healthy volunteers was analyzed using the same methods and is presented as the normal range (median ±range). Serum concentrations of (**A**) IL-6, (**B**) TNF-α, (**C**) IL-1β, (**D**) IL-18, and (**E**) IFN-γ are presented. Patients 01–08 received one dose of Allocetra-OTS on day 1 and patients 09–12 received two doses on days 1 and 3.

**Figure 5B.**
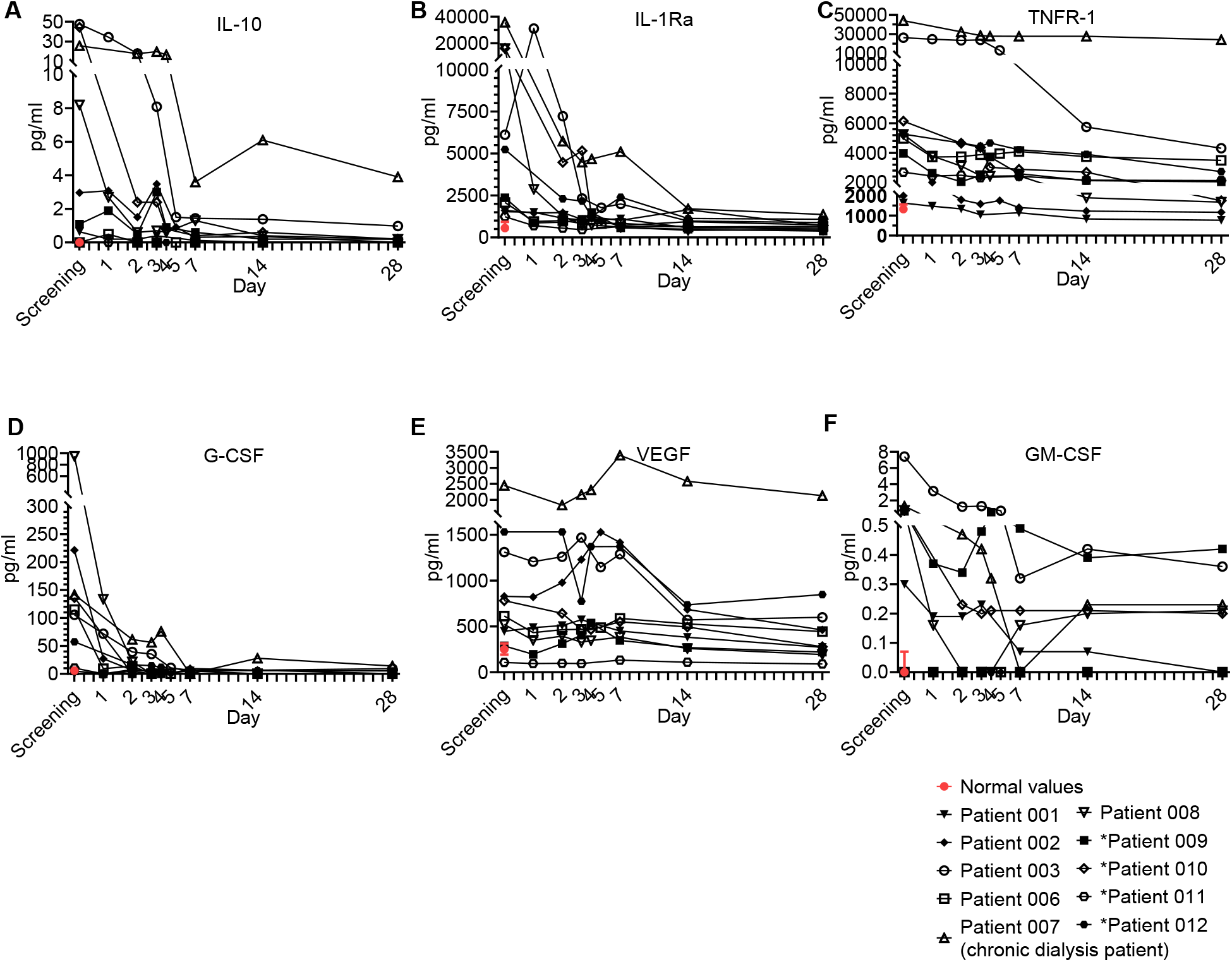
Anti-Inflammatory cytokine and growth factor kinetics during sepsis. Serum was obtained from patients at the indicated times and cytokine/growth factor analysis was performed as described in Methods. The serum of three healthy volunteers was analyzed using the same methods and is presented as the normal range (median ± range). Serum concentrations of (**A**) IL-10, (**B**) IL-1Ra, (**C**) TNF-R1, are presented in the upper panel. (**D**) G-CSF, (**E**) VEGF, and **(F)** GM-CSF, are presented in the lower panel. Patients 01–08 received one dose of Allocetra-OTS on day 1 and patients 09–12 received two doses on days 1 and 3.

**Figure 5C.**
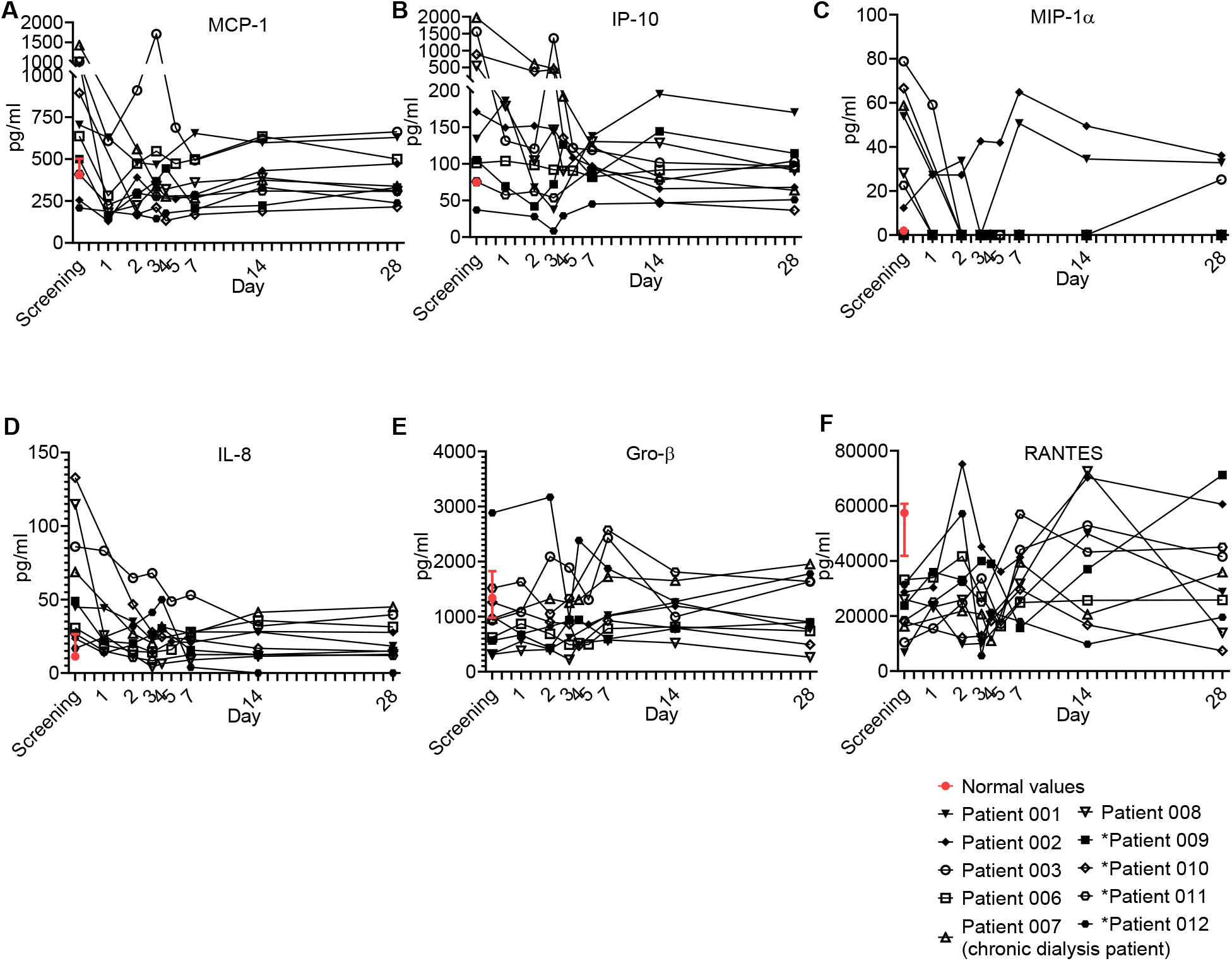
Chemokine kinetics during sepsis. Serum was obtained from patients at the indicated times and cytokine/growth factors analysis was performed as described in Methods. The serum of three healthy volunteers was analyzed using the same methods and is presented as the normal range (median ± range). Serum concentrations of (**A**) MCP-1, (**B**) IP-10, (**C**) MIP-1α, (**D**) IL-8, (**E)** GRO-?, and **(F)** RANTES, are presented. Patients 01–08 received one dose of Allocetra-OTS on day 1 and patients 09–12 received two doses on days 1 and 3.

**Figure 5D.**
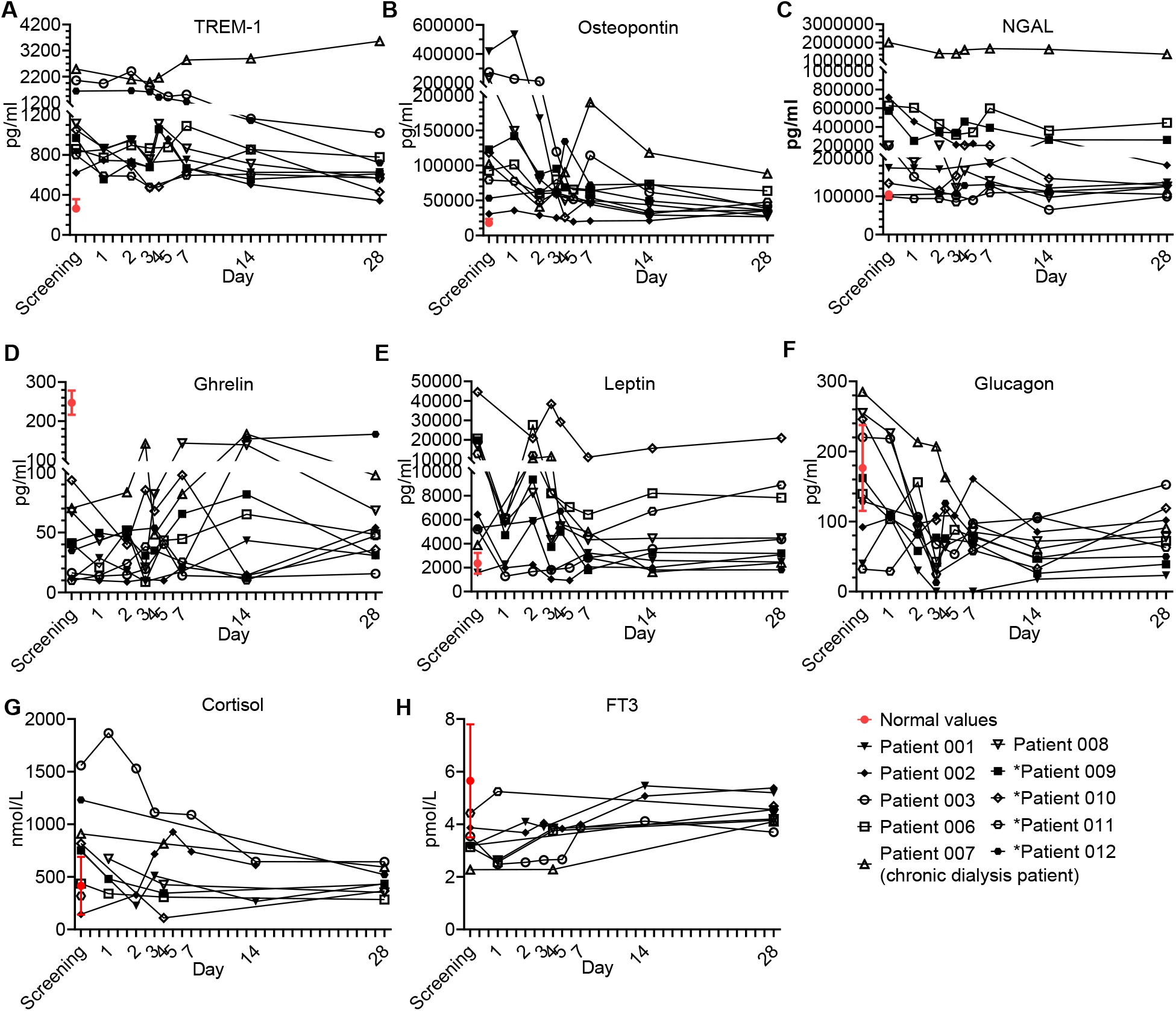
Immune modulator and endocrine hormone kinetics during sepsis. Serum was obtained from patients at the indicated times and cytokine/growth factors analysis was performed as described in Methods. The serum of three healthy volunteers was analyzed using the same methods and is presented as the normal range (median ± range). Serum concentrations of **(A)** TREM-1, **(B)** osteopontin, **(C)** N-GAL, **(D)** ghrelin, **(E)** leptin, and **(F)** glucagon, **(G)** cortisol, and **(H)** FT3, are presented in the lower panel. Patients 01–08 received one dose of Allocetra-OTS on day 1 and patients 09–12 received two doses on days 1 and 3.

**Figure 5E.**
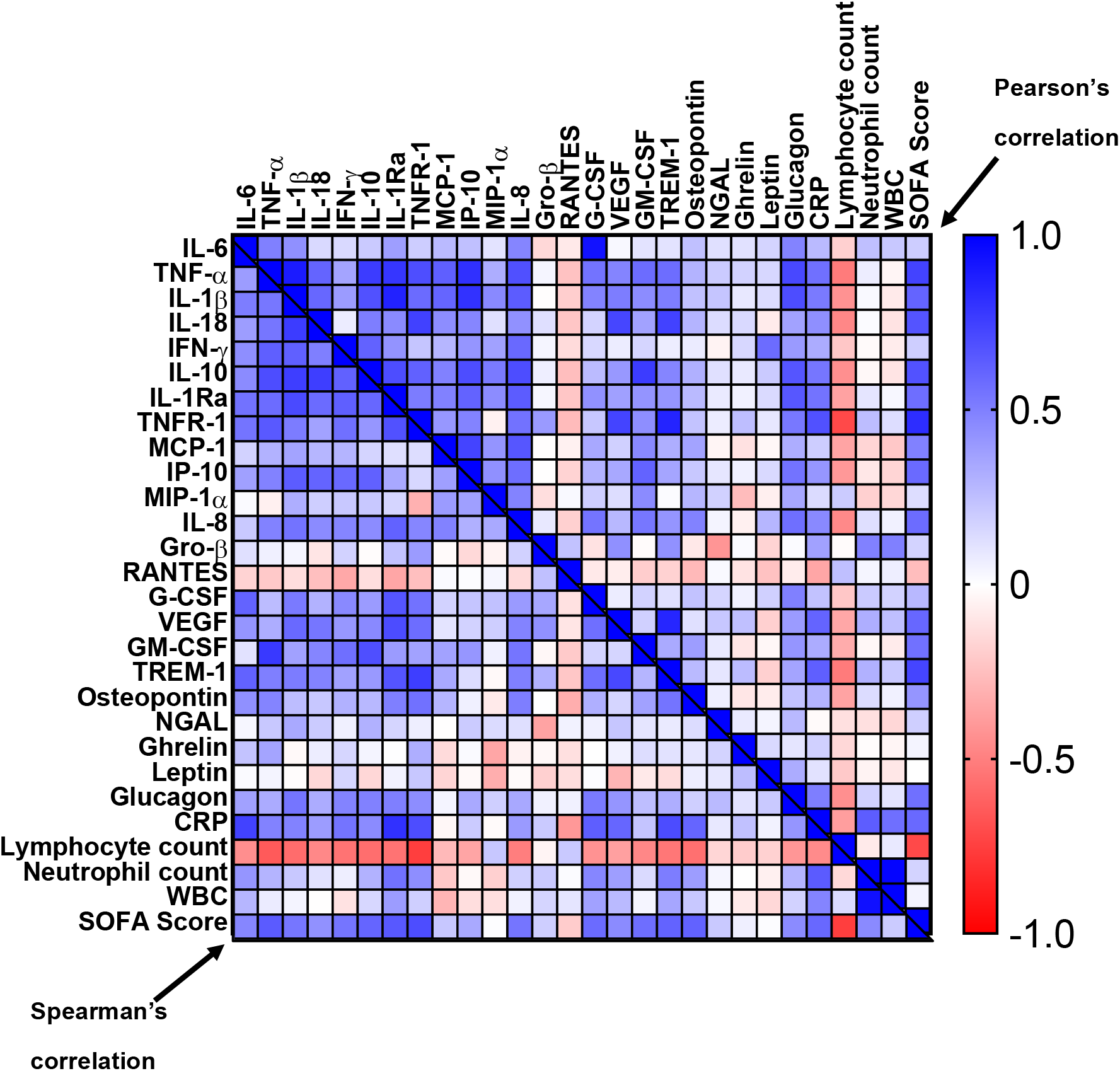
Correlation matrix of cytokines, chemokines, and immunomodulators with SOFA score, CRP, and blood counts. A correlation heat-map of the tested cytokines/chemokines, HGFs, and immunomodulators with CRP, SOFA score, and blood counts. Pearson’s r values are on the top right and the Spearman’s ρ values are on the bottom left.

Overall, all patients had elevated TNF-a and IL-6 levels at initial screening and most had elevated IL-1β and IL-18, while some had elevated IFN-γ; however, a gradual decrease for levels of all of the cytokines studies was observed upon resolution of sepsis. These trends of pro-inflammatory cytokines were characteristic to most of the patients.

### Anti-inflammatory cytokines

Interestingly, anti-inflammatory cytokines are also upregulated in sepsis in parallel to pro-inflammatory cytokines (Figure 5B) and were suggested to have a late contribution to sepsis-related immunosuppression.^24^ Four anti-inflammatory cytokines were tested, including IL-10, IL-1Ra, IL-27, and soluble TNFR-1. IL-27 was not detected in our patients, and the other anti-inflammatory cytokines are presented in Figure 5B, upper panel. Overall, the trend of the anti-inflammatory cytokines resembled that of the pro-inflammatory cytokines. Most patients had elevated IL-10, IL-1Ra, and TNFR-1, which gradually decreased as sepsis resolved.

### Hematopoietic growth factors (HGFs)

During sepsis, immune cells undergo profound phenotypic modifications in their activation state, response to stimuli, and localization. These phenomena are finely regulated by various cytokines and HGFs. An HGF is a relatively stable, secreted, or membrane-bound glycoprotein that causes immune cells to mature and/or proliferate. They also have profound effects on cell functions and behaviors. HGFs are deeply involved in sepsis pathophysiology both in the initial and the late phases.^25^ Four growth factors were tested in the study, including G-CSF, VEGF, GM-CSF (Figure 5B) and LIF (a growth factor-like cytokine), which was undetected. Overall, the trend of the HGFs resembled that of the pro- and anti-inflammatory cytokines and chemokines. Most patients presented with elevated G-CSF, VEGF, and to a lesser extent, GM-CSF at initial screening; for all, a gradual decrease upon resolution of sepsis was observed. These trends of HGFs were characteristic of most patients that received a single dose of Allocetra-OTS.

### Chemokines

Chemokines play pivotal roles in regulating the migration and infiltration of monocytes/macrophages and neutrophils to sites of inflammation, and as such, they are usually involved in sepsis.^26^ Six chemokines were tested and are presented here, including MCP-1, IP-10, MIP-1α, IL-8, Gro-β, and RANTES. Overall, most patients were screened with elevated MCP-1, IP-10, MIP-1α and IL-8; for all, a gradual decrease upon resolution of sepsis was observed (Figure 5C). Of exception were Gro-β and RANTES, which were detected in most patients below the normal levels and increased upon resolution of sepsis. These trends of chemokines were characteristic to most of the patients that received a single dose of Allocetra-OTS.

### Other immuno-modulating agents

Several miscellaneous immuno-modulating factors were also tested as part of the immune modulating effect of Allocetra-OTS on the resolution of sepsis (Figure 5D). These factors included TREM-1, osteopontin, NGAL, gherlin, leptin, and several endocrinological hormones. Overall, all the patients presented with elevated TREM-1 and OPN and most had elevated NGAL and leptin levels; for all, a gradual decrease upon resolution of sepsis was observed. Of exception is ghrelin, which was below the normal levels in all the patients at presentation and increased upon resolution of sepsis. These trends of immuno-modulating factors were characteristic of most of the patients that received a single dose of Allocetra-OTS.

### Endocrinology kinetics

Five hormones were tested in all subjects: Cortisol, FT3, FT4, Glucagon and Insulin. Additional hormones (TSH, ACTH and GH) were tested in some subjects. Sepsis is considered an acute stress response with a release of stress hormones including cortisol and glucagon.^27^ There were elevated cortisol levels in 7/10 patients at presentation, with the average of all the patients being above normal (752 nmol/L, normal range of 140–690 nmol/L). Patients 03 and 12 (both with pneumonia) had the highest levels of cortisol at screening. Following infusion of the Allocetra-OTS, cortisol levels were downregulated (including Patient 03), reaching normal concentrations by Day 28 (Figure 5D). Interestingly, serum glucagon had similar kinetics to cortisol, and 7/10 patients had high glucagon at screening (average level of all patients = 150pg/ml, range 32–285 pg/ml) compared to normal (average of 73 pg/ml, range 30–115 pg/ml), with a rapid decline in the first 2–3 days reaching low levels on day 28. Insulin levels were in the low normal range upon presentation (opposite to glucagon) and the levels were normalized upon resolution of sepsis. ACTH and GH were tested in some subjects and were within the normal range throughout the study, except for patient 07 (chronic dialysis) who had elevated ACTH levels on day 28 (21·5 pMol/L versus normal levels up to 13·5). All patients were screened for FT3 and FT4 levels and all had low normal or below normal FT3 levels with gradual increase thereafter as sepsis resolved, but normal FT4 and TSH levels at presentation and during the resolution of sepsis (Figure 5D). These changes in serum thyroid function associated with acute illness have been termed ‘euthyroid sick syndrome’ or ‘low T3 syndrome’. FT3 production during the acute stress response in sepsis is inhibited by both cortisol and IL-6.^28-30^ We therefore tested whether the recovery of FT3 seen in the patients was linked to the downregulation of IL-6 and/or cortisol. Indeed, a strong negative correlation was found between FT3 and IL-6 (ρ-Spearman= -0·73) and an intermediate negative correlation was found between FT3 and cortisol and glucagon (ρ-Spearman= -0·54, -0·56, respectively). The euthyroid sick syndrome should thus not be viewed as an isolated pathologic event but as part of a coordinated systemic reaction to sepsis involving both the immune and endocrine systems.

In summary, most patients had elevated pro-, and anti-inflammatory cytokines/chemokines/immune modulators/stress factors, with a gradual normalization upon resolution of sepsis (Figure 5). The physiological responses to acute stress in sepsis results from the activation of an array of factors, including immunological, neural, and hormonal pathways that pose a threat to the homeostasis of the organism and elicit a common series of adaptive physiological responses. We created a heat map (Figure 5E), correlating the different immunological components of the response to stress to blood counts and SOFA score. As shown, high correlation is seen between pro- and anti-inflammatory cytokines, chemokines, HGFs with WBC counts, neutrophils, CRP, and most importantly with SOFA score. Inverse characteristic correlation is shown with lymphocyte counts, and the chemokines Gro-β and RANTES.

## Discussion

Apoptotic cells have immunomodulatory functions via their interaction with macrophages and dendritic cells, and their administration was suggested to be used as a potential therapeutic intervention.^13,14,31-33^ Allocetra-OTS preparation is unique, with an emphasis on early apoptosis (i.e. apoptotic cells are Annexin V^+^ propidium iodide^−^) to avoid any effect from necrotic cells. Although not all the molecular events underlying the potential immune-regulating function of apoptotic cells are clear, changes in macrophages and dendritic cells towards a homeostatic phenotype have been investigated by several authors and implicated in apoptotic cell-mediated immune modulation (reviewed by^13,32,33^). Local administration of apoptotic cells has been used to attenuate both bleomycin- and lipopolysaccharide (LPS)-induced lung inflammation, with reduced neutrophil recruitment into the lung, enhanced phagocytosis by alveolar macrophages, and reduced pro-inflammatory cytokine production.^34^ Infusion of apoptotic cells 24 hours after initiation of sepsis has also been shown to protect against mortality in a mouse model of sepsis, with reduced pro-inflammatory cytokine and neutrophil recruitment into organs.^35^

Many of the measured cytokines/chemokines/immune-modulators were proven to be pathogenic in sepsis and septic shock; however, single targeting of cytokines did not ameliorate sepsis in many trials using anti-TNF, anti-IL-1β, and other cytokines in sepsis.^36^

Interestingly, and as suggested elsewhere,^37^ anti-inflammatory cytokines are also elevated early in the course of sepsis. This supported the notion that secretion of pro- and anti-inflammatory mediators in septic shock occurs as a simultaneous immune response program initiated early in the course of the disease,^38^ and in severe sepsis, the IL-10/lymphocyte ratio was significantly correlated with the APACHE II score and strongly predicted 28-day mortality.^39^ Apoptotic cell infusion represents a more holistic approach, leading to rebalancing of all pro- and anti-inflammatory cytokines and chemokines, growth factors, and other immuno-modulating agents, and reprogramming of monocytes/macrophages and dendritic cells.^13^

In the first few days of clinically apparent infection, there is an innate immune response in all patients. Gene expression studies have shown that transcripts from sepsis and severe viral infections involve pathways associated with signaling through TLRs, IL-27, IL-12, IFN-γ, type I IFN-inducible transcripts, and the JAK-STAT pathway that are overabundant in the acute phase and remain elevated until defervescence occurs and may lead to endothelial dysfunction.^40^ The mechanism through which cytokines such as TNF might mediate endothelial dysfunction are not clear, though changes in the integrity of inter-endothelial cell junctions is a possible cause. Most permeability-inducing factors bind to endothelial cell plasma membrane receptors, activate heterotrimeric G proteins, and cause an increase in intracellular Ca2^+^. This results in myosin-driven endothelial contraction and opening of tight junctions. In some viral diseases, increased capillary permeability occurs when viremia is in steep decline and serum cytokine concentrations are at or near their peak levels.^40^ In addition, pathogen-induced lung injury can progress into acute lung injury or its more severe form, acute respiratory distress syndrome (ARDS), as seen with sepsis and SARS-CoV or influenza virus infections.

To further evaluate apoptotic cell effect a randomized-controlled trial is needed, but these results may reflect a novel and safe mechanism for treatment of sepsis as well as diseases associated with cytokine storm such as flu complications, acute lung injury and acute respiratory distress syndrome as seen in SARS-CoV and influenza virus infections,^41^ CAR-T therapy,^42,43^ and the recently described COVID-19.^44,45^

## Data Availability

The data that support the findings of this study are available from the corresponding author, upon reasonable request

## Author contributions

PVH: performed the study and wrote the manuscript; AA performed the study and participated in writing; SS performed the study and participated in writing; YH prepared the historical control section; EZ: performed the study and collected data; AN: performed the study and collected data; SZ performed the study and collected data, RA analyzed blood samples; YS analyzed blood samples; BR prepared figures and participated in writing; BF performed the study and collected data; DM designed the study and wrote the manuscript.

## Notes

### Clinical Trial

NCT03925857

### Author Declarations

The study was approved by the Human Subjects Review Board of The Helsinki Committee, Hadassah Medical Organisation, Jerusalem, Israel. Approval number: HMO-0152-18.

